# Dopamine pathway and Parkinson’s risk variants are associated with levodopa-induced dyskinesia

**DOI:** 10.1101/2023.08.28.23294610

**Authors:** Yuri L. Sosero, Sara Bandres-Ciga, Bart Ferwerda, Maria T. P. Tocino, Dìaz R. Belloso, Pilar Gómez-Garre, Johann Faouzi, Pille Taba, Lukas Pavelka, Tainà M. Marques, Clarissa P. C. Gomes, Alexey Kolodkin, Patrick May, Lukasz M Milanowski, Zbigniew K. Wszolek, Ryan J. Uitti, Peter Heutink, Jacobus J. van Hilten, David K. Simon, Shirley Eberly, Ignacio Alvarez, Lynne Krohn, Eric Yu, Kathryn Freeman, Uladzislau Rudakou, Jennifer A. Ruskey, Farnaz Asayesh, Manuel Menéndez-Gonzàlez, Pau Pastor, Owen A. Ross, Rejko Krüger, the NCER-PD Consortium, Jean-Christophe Corvol, Sulev Koks, Pablo Mir, Rob M.A. De Bie, Hirotaka Iwaki, Ziv Gan-Or, the International Parkinson’s Disease Genomic Consortium

## Abstract

**Background:** Levodopa-induced dyskinesia (LID) is a common adverse effect of levodopa, one of the main therapeutics used to treat the motor symptoms of Parkinson’s disease (PD). Previous evidence suggests a connection between LID and a disruption of the dopaminergic system as well as genes implicated in PD, including *GBA1* and *LRRK2*.

**Objectives:** To investigate the effects of genetic variants on risk and time to LID.

**Methods:** We performed a genome-wide association study (GWAS) and analyses focused on *GBA1* and *LRRK2* variants. We also calculated polygenic risk scores including risk variants for PD and variants in genes involved in the dopaminergic transmission pathway. To test the influence of genetics on LID risk we used logistic regression, and to examine its impact on time to LID we performed Cox regression including 1,612 PD patients with and 3,175 without LID.

**Results:** We found that *GBA1* variants were associated with LID risk (OR=1.65, 95% CI=1.21-2.26, p=0.0017) and *LRRK2* variants with reduced time to LID onset (HR=1.42, 95% CI=1.09-1.84, p=0.0098). The fourth quartile of the PD PRS was associated with increased LID risk (OR_fourth_quartile_=1.27, 95% CI=1.03-1.56, *p=*0.0210). The third and fourth dopamine pathway PRS quartiles were associated with a reduced time to development of LID (HR_third_quartile=_1.38, 95% CI=1.07-1.79, *p=*0.0128; HR_fourth_quartile=_1.38, 95% CI=1.06-1.78, *p=*0.0147).

**Conclusions:** This study suggests that variants implicated in PD and in the dopaminergic transmission pathway play a role in the risk/time to develop LID. Further studies will be necessary to examine how these findings can inform clinical care.

## Introduction

Levodopa is one of the most commonly administered therapies for Parkinson’s disease (PD), particularly to treat motor symptoms.^1^ However, as the disease progresses and patients are exposed to long-term levodopa therapy, a significant proportion develops levodopa-induced dyskinesia (LID), a debilitating side effect characterized by involuntary, uncontrolled, and often choreiform movements.^2^ LID is estimated to affect around 40-50% of PD patients within 4-6 years of initiating levodopa therapy,^3, 4^ however, a subset of them manifests LID also within the first year of the therapy,^5^ demonstrating the broad variability of LID risk and onset. The most widely accepted pathophysiologic hypothesis suggests that LID development is connected with a pulsatile stimulation of the dopamine receptors in the nucleus striatum.^6^ This phenomenon occurs due to the progressive dopaminergic loss in PD, resulting in impaired presynaptic storage capacity of dopamine, and is exacerbated by elevated doses of levodopa.^6–8^ Other pathways have also been implicated in LID development, including the glutamatergic, serotonergic and noradrenergic neural circuits.^7, 8^

Multiple environmental risk factors affecting LID have been identified, including levodopa dosage and duration of the therapy, use of dopamine agonists, PD age at onset (AAO), disease duration and severity, female sex and lower body mass index (BMI).^9–13^ Most of the suggested genetic risk factors for LID are related to the dopamine pathway, including genes encoding the dopamine receptors, especially *DRD2* and *DRD3*,^14–16^ the dopamine transporter *SLC6A3*,^17, 18^ or enzymes that metabolize dopamine and are targeted by PD therapeutics,^19, 20^ catechol-O-methyltransferase (*COMT*)^21–23^ and monoamine oxidases A and B (*MAOA*, *MAOB*).^22–24^ Interestingly, variants in *GBA1* and *LRRK2,* among the most frequent genetic risk factors for PD,^25, 26^ have also been identified as potential risk factors for LID.^27–32^ Carriers of *GBA1* and *LRRK2* variants show distinctive clinical presentations in PD, with *GBA1* variants being associated with a more rapidly progressive PD with earlier onset,^33^ and *LRRK2* variants with an overall more benign disease course, but with also more frequent postural instability and gait difficulty as well as slightly earlier AAO compared to sporadic PD.^34^ Other variants reported in LID include those in *BDNF,* involved in neural plasticity,^35, 36^ *GRIN2A,* encoding a glutamatergic receptor,^37^ and *ADORA2A*, encoding the adenosine A2a receptor gene.^38^ However, the association between LID and most of the above-mentioned putative genetic risk factors is still controversial, with most findings reported deriving from candidate genes studies that failed to be confirmed in replication studies. ^39–44^

Here, we aimed to systematically evaluate how genetics affect the risk and rate of progression to LID including a total of 4,787 PD patients from multiple centers. For this purpose, we performed genome-wide association studies (GWAS) and downstream analyses focused on specific genes previously implicated in LID. In addition, we tested the effect produced by cumulative genetic risk on the occurrence and rate of progression to LID, including risk variants previously associated with PD and variants in genes involved in the dopaminergic transmission pathway.

## Methods

### Population

The study population included a total of 4,787 PD patients, of which 1,612 with and 3,175 without LID (Table 1). PD was diagnosed by movement disorder specialists according to the UK Brain Bank or International Parkinson Disease and Movement Disorders Society criteria.^45^ LID diagnosis was made based on the Unified Parkinson’s Disease Rating Scale (UPDRS) part IV and direct clinical evaluation. The participants were of European ancestry and their clinical and genetic data were collected from 15 different cohorts (Table 1), 12 of which were from the International Parkinson’s Disease Genomics Consortium (IPDGC) and 3 from the Accelerating Medicines Partnership Parkinson’s Disease (AMP-PD, https://amp-pd.org/). The latter includes the Parkinson’s Disease Biomarkers Program (PDBP), Parkinson’s Progression Markers Initiative (PPMI) and Harvard Biomarker Study (HBS) cohorts. The cohorts were included in the different analyses depending on data availability. The cohorts included in each analysis are specified in Supplementary Table 1.

**Table 1.** Demographic characteristics of PD patients in the individual cohorts.

| **Center** | **LID-,**<br>**n** | **Age LID-**<br>**(SD)** | **%Mal**<br>**LID-** | **LID+,**<br>**n** | **Age LID+**<br>**(SD)** | **%Mal**<br>**LID+** | **Tot** |
| --- | --- | --- | --- | --- | --- | --- | --- |
| **Barcelona** | 103 | 73.3 (10.9) | 50% | 48 | 72.3 (7.7) | 40% | 151 |
| **CORIELL** | 221 | 62.7 (8.9) | 67% | 117 | 61.7 (9.6) | 59% | 338 |
| **DIGPD** | 220 | 67.5 (9.3) | 62% | 166 | 63.9 (10.4) | 56% | 386 |
| **LEAP** | 336 | 68.9 (8.8) | 75% | 75 | 67.7 (8.5) | 50% | 411 |
| **Luxembourg** | 330 | 67.8 (11.4) | 66% | 140 | 66.2 (10.0) | 66% | 470 |
| **Mayo Clinic Florida** | 404 | 75.8 (9.9) | 69% | 151 | 72.0 (10.1) | 62% | 555 |
| **McGill** | 258 | 63.2 (16.5) | 34% | 120 | 61.3 (15.7) | 43% | 378 |
| **Oviedo-HUCA** | 80 | 69.8 (8.9) | 51% | 110 | 70.4 (10.5) | 55% | 190 |
| **PDBP – PPMI – HBS**<br>**(AMP-PD)** | 580 | 58.4 (12.4) | 66% | 87 | 56.1 (12.2) | 53% | 667 |
| **PreCEPT** | 181 | 61.6 (8.6) | 68% | 137 | 58.5 (9.7) | 66% | 318 |
| **SCOPA** | 109 | 59.1 (10.9) | 66% | 177 | 60.0 (10.6) | 62% | 286 |
| **Sevilla** | 180 | 69.7 (10.9) | 61% | 252 | 66.0 (11.1) | 57% | 432 |
| **Tartu** | 173 | 73.0 (8.2) | 38% | 32 | 72.0 (8.6) | 50% | 205 |
| **TOTAL** | 3175 | 67.0 (10.4) | 52% | 1612 | 65.2 (10.4) | 54% | 4787 |

AMP-PD: Accelerating Medicines Partnership Parkinson's disease, including the Parkinson's Disease Biomarkers Program (PDBP), Parkinson's Progression Markers Initiative (PPMI) and Harvard Biomarker Study (HBS) cohorts; Barcelona: Hospital Universitari Mutua de Terrassa, Spain; CORIELL: NINDS Exploratory Trials in PD Long-Term Study 1 (NET-PD LS1), Coriell Institute for Medical Research, USA; DIGPD: Drug Interaction With Genes in Parkinson's Disease, France; LEAP: Levodopa in Early Parkinson's Disease, Netherlands; Luxembourg: Luxembourg Centre for Systems Biomedicine; Mayo Clinic Florida: Mayo Clinic Florida, USA; McGill: McGill University, Canada; Oviedo: Central University Hospital of Asturias, Spain; PreCEPT: Parkinson Research Examination of CEP-1347 Trial; SCOPA: Scales for Outcomes in Parkinson's disease; Sevilla: Universidad de Sevilla; Tartu: University of Tartu; LID-, n: individuals without levodopa-induced dyskinesia; LID+, n: individuals with levodopa-induced dyskinesia; Age (SD): mean age (standard deviation); %Mal: percentage of males; Tot: total number of individuals per cohort.

### Genetic analyses

Excluding the AMP-PD cohorts, with whole genome sequencing (WGS) data, the other centers (Table 1) were genotyped using the OmniExpress, NeuroX,^46^ NeuroChip^47^ or MegaChip GWAS array according to the manufacturer’s instructions (Illumina Inc.). Quality control was performed following standard pipelines (detailed in https://github.com/neurogenetics/GWAS-pipeline) using Plink 1.9.^48^ In brief, we filtered out heterozygosity outliers using an F-statistic cut-off of <-0.15 or >0.15. Individuals with a variant call rate <95% and sex mismatch were excluded. Variants missing in >5% of the participants, with disparate missingness between cases and controls (*p*<1E-04), or significantly deviating from Hardy-Weinberg equilibrium in controls (p<1E-04) were also removed. We used GCTA to check for relatedness closer than first cousins between participants (PIHAT>0.125). We performed imputation using the Michigan imputation server (https://imputationserver.sph.umich.edu/index.html<u>#</u>) with the Haplotype Reference Consortium reference panel r1.1 2016 under default settings. Ancestry outliers were detected using HapMap3 principal component analysis (PCA) data in R version 4.0.1.

After imputation, we selected variants with r2>0.8 and a minor allele frequency (MAF)>0.05, while retaining common risk variants in the *GBA1* (p.N370S, p.E326K and p.T369M) and *LRRK2* (p.G2019S, p.M1646T and p.R1441G/C) regions, to perform specific analyses on these variants (detailed below). These genes were specifically selected given their importance in PD etiology^25, 26^ and recent clinical trials^49^ as well as their previously suggested association with LID.^27–32^ The carrier status of *GBA1/LRRK2* risk variants in individuals with and without LID is detailed in Supplementary Table 2 and Supplementary Table 3. Carriers of variants in the same gene were combined, so that the carrier status for *GBA1* and *LRRK2* refers to any aforementioned *GBA1* and *LRRK2* variants, respectively. To examine the association between the *GBA1* and *LRRK2* risk variant carrier status and LID occurrence we performed logistic regression, and to evaluate the association between the carrier status and time to LID onset we performed Cox regression using the R package “survival” (https://cran.r-project.org/web/packages/survival/). The time to LID variable included in the Cox regression was defined as the period between the start of levodopa therapy and LID onset, as previously done.^50^ When LID did not manifest, this parameter was right-censored at the last follow-up. We adjusted the analyses by multiple covariates including principal components (PCs), PD AAO, sex, levodopa dosage, levodopa equivalent daily dose (LEDD),^51, 52^ dopamine agonist use, BMI, Hoehn and Yahr score (HY) and, exclusively for logistic regression, disease duration. For logistic regression analyses, we included the cumulative levodopa dosage and LEDD starting from the baseline (i.e., levodopa initiation) to the last time point (i.e., LID onset or last follow-up when LID was not present). In Cox regression, to avoid collinearity with the time to LID onset dependent variable, we replaced cumulative doses with doses at the last time point. All the covariates were selected using an Akaike Information Criterion (AIC)-based stepwise regression approach, which evaluated the model goodness of fit and selected the most appropriate covariates to include in the model. We performed the analyses separately in each cohort and then meta-analyzed the results using the R package “metafor” (https://cran.r-project.org/web/packages/metafor/index.html). Since variants in these genes have been previously associated with LID, we used a significance threshold of α=0.05.

Similar to the analyses on specific genes, to investigate the overall impact of genetics on LID risk and time to onset we also performed GWAS with, respectively, logistic and Cox regression adjusted for the above-specified covariates. Cox regression was performed using the SurvivalGWAS_SV software (https://www.liverpool.ac.uk/population-health/research/groups/statistical-genetics/survival-gwas-sv/).^53^ We conducted the analyses in each cohort separately, and then meta-analyzed the results using the METAL software (https://genome.sph.umich.edu/wiki/METAL_Documentation) with a fixed effects model weighted by β coefficients and the inverse of the standard errors.

### PD risk variant-based polygenic risk score

To assess the impact on LID of the cumulative genetic risk for PD we calculated polygenic risk score (PRS) for each PD patient including the 90 variants associated with PD in the most recent GWAS meta-analysis in Europeans.^54^ PRS calculation was performed based on the weighted allele dose as implemented in PRSice2 using default clumping (https://choishingwan.github.io/PRSice/).^55^ To investigate the association between the PRS and LID risk we performed logistic regression, while to evaluate the association between PRS and progression to LID we performed Cox regression. The analyses were adjusted for PCs, PD AAO, sex, HY and levodopa dosage, cumulative in logistic regression and at the last time point in Cox regression. These analyses were repeated using PRS as a continuous variable and then as a discrete variable by dividing the PRS into quartiles. For the analysis using PRS quartiles, we separately compared the association of individual membership to the second, third and fourth quartiles vs the first quartile with LID risk/progression.

### Dopamine pathway polygenic risk score

To assess the impact of genes involved in the dopaminergic transmission pathway we also constructed a pathway polygenic risk score, or polygenic effect score (PES)^56^ using the PRSet feature of PRSice2 (https://choishingwan.github.io/PRSice/prset_detail/). Genes involved in this pathway were obtained from Explore the Molecular Signatures Database (MSigDB, version 2023.1), a collection of annotated gene sets for use with Gene Set Enrichment Analysis (GSEA) software (https://www.gsea-msigdb.org/gsea/msigdb/). These genes included *CDK5, FLOT1, PARK7, CHRNB2, ADORA2A, CRH, CRHBP, DRD1, DRD2, DRD3, DRD4, DRD5, TOR1A, RASD2, PNKD, GDNF, ARRB2, PRKN, PTGS2, RAB3B, PINK1, SLC6A2, SLC, 6A3, SLC6A4, SNCA, TH, CNTNAP4* (detailed at http://www.gsea-msigdb.org/gsea/msigdb/human/geneset/GOBP_SYNAPTIC_TRANSMISSION_DOPAMINERGIC). To select the variants in each of those genes to include in the analyses we used the LID GWAS meta- analysis summary statistics, filtering variants with a p-value less than or equal to 0.05. In addition, we performed linkage disequilibrium (LD) clumping using the default r2=0.1 and selecting variants at 250 Kb of distance from the pathway-related genes. A total of 1000 permutations were implemented to generate the empirical p-value corresponding to the optimized PES prediction of the dependent variable in the target cohort. We then calculated individual PES for each target cohort. To avoid potential inflation due to the presence of the target cohort in the meta-analysis summary statistics, each time we calculated the PES for a target cohort we excluded such cohort from the meta-analysis using a leave-one-out approach. To investigate the association between the dopamine pathway PES and LID risk we performed logistic regression, while to evaluate the association between the PES and progression to LID we performed Cox regression, as specified above for the PRS analyses.

## Results

### GBA1 and LRRK2 variants show significant associations with LID risk and time to LID

Analyses focusing on *GBA1* showed that *GBA1* variants were significantly associated with LID risk (OR=1.65, 95% CI=1.21-2.26, *p*=0.0017, Fig. 1A). No association was found with time to LID (HR=1.25, 95% CI=0.99-1.58, *p*=0.0635, Fig. 1B). In contrast, *LRRK2* variants showed no association with LID risk (OR=1.18, 95% CI=0.84-1.67, *p*=0.3484, Fig. 2A) but were significantly associated with reduced time to development of LID (HR=1.42, 95% CI=1.09-1.84, *p*=0.0098, Fig. 2B)

**Fig. 1.**
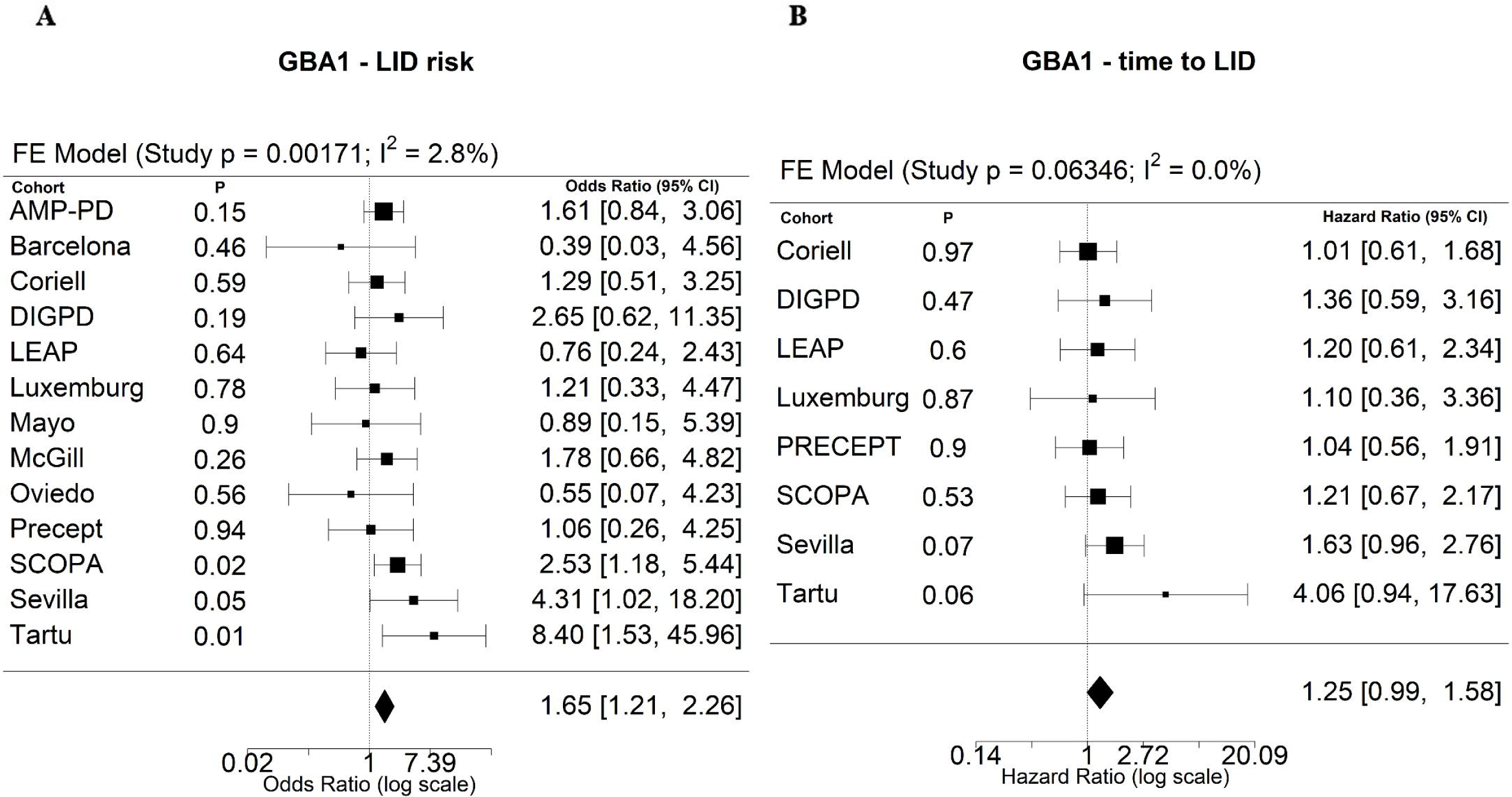
A-B – Association between *GBA1* variants and LID. The meta-analysis forest plot shows the coefficient (black squares) and 95% confidence interval (bars) of the analyses in each single cohort. The size of the square is proportional to the weight the cohort had on the overall meta-analysis, based on their single standard error. The black diamond at the bottom represents the overall coefficient and confidence interval. **A**. Logistic regression between *GBA1* variants and LID risk; **B**. Cox regression between *GBA1* variants and time to development of LID. FE: fixed effect model; AMP-PD: Accelerating Medicines Partnership Parkinson’s disease, including the New Discovery of Biomarkers (BioFIND), the Harvard Biomarker Study (HBS) and the Parkinson’s Disease Biomarkers Program (PDBP) cohorts; Barcelona: Hospital Universitari Mutua de Terrassa, Spain; CORIELL: NINDS Exploratory Trials in PD Long-Term Study 1 (NET-PD LS1), Coriell Institute for Medical Research, USA; DIGPD: Drug Interaction With Genes in Parkinson’s Disease, France; LEAP: Levodopa in Early Parkinson’s Disease, Netherlands; Luxemburg: Luxembourg Centre for Systems Biomedicine; Mayo: Mayo Clinic, USA; McGill: McGill University, Canada; Oviedo: Central University Hospital of Asturias, Spain; PreCEPT: Parkinson Research Examination of CEP-1347 Trial; SCOPA: SCales for Outcomes in PArkinson’s disease; Sevilla: Universidad de Sevilla; Tartu: University of Tartu

**Fig. 2.**
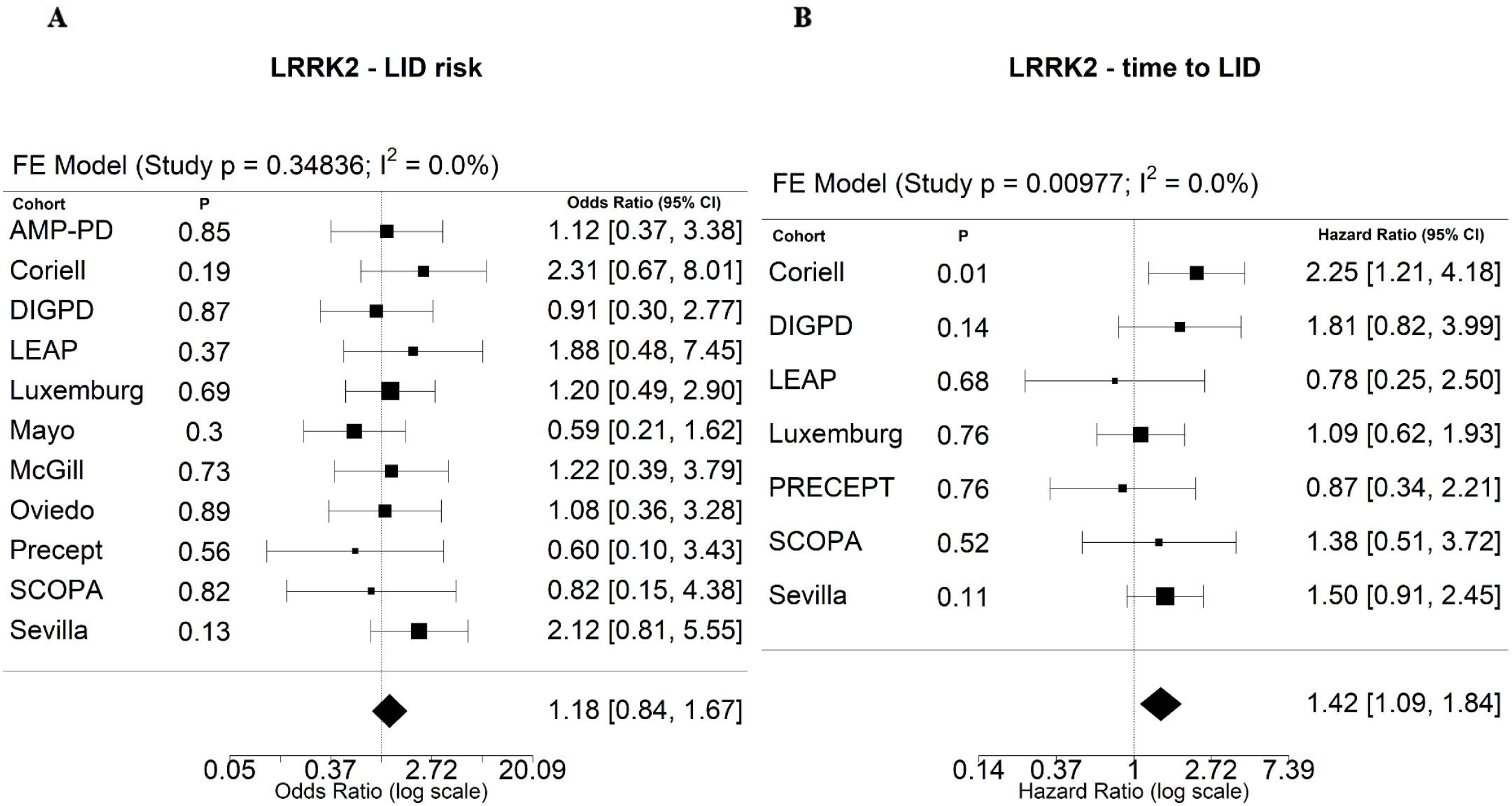
A-B - Association between *LRRK2* variants and LID. **A**. Logistic regression between *LRRK2* variants and LID risk; **B**. Cox regression between *LRRK2* variants and time to development of LID.

In the GWAS genomic inflation was evaluated using quantile-quantile plots (Q-Q plots) and the lambda factor, showing no inflation and a slight deflation (lambda logistic regression=0.9709, lambda Cox regression=0.9555, Supplementary Fig. 1-2). GWAS using both logistic and Cox regression showed no significant association with LID risk or time to development of LID, respectively (Supplementary Fig. 3, Supplementary Fig. 4). We further examined whether variants previously associated with LID in the literature^14–18, 21–24^ and from the LIDPD website (http://LiDpd.eurac.edu/) showed associations in the current GWAS, but we found no significant results (Supplementary Tables 4-5). A recent GWAS in LID (Martinez et al., 2023, MedRxiv) nominated significant signals in a progression GWAS meta-analysis. However, our study failed to confirm these findings and the reported variants did not reach the nominal significance of 0.05 in our GWAS (Supplementary Table 6).

### PD risk variant-based polygenic risk score is associated with increased risk for LID

PRS analyses aggregating PD-associated variants showed that higher values of PRS were associated with a very mild increase in LID risk (OR=1.02, %95 CI=1.002-1.035, *p=*0.0298, Fig. 3B). When dividing the PRS in quartiles, logistic regression showed a significant association between the fourth quartile and LID, with a greater risk compared to the analyses using PRS as a continuous variable (OR_fourth_quartile_=1.27, 95% CI=1.03-1.56, *p=*0.0210, Fig. 3A, Supplementary Table 7). Cox regression did not show any significant associations between PRS and time to development of LID (Supplementary Fig. 5 A-B, Supplementary Table 8).

**Fig. 3.**
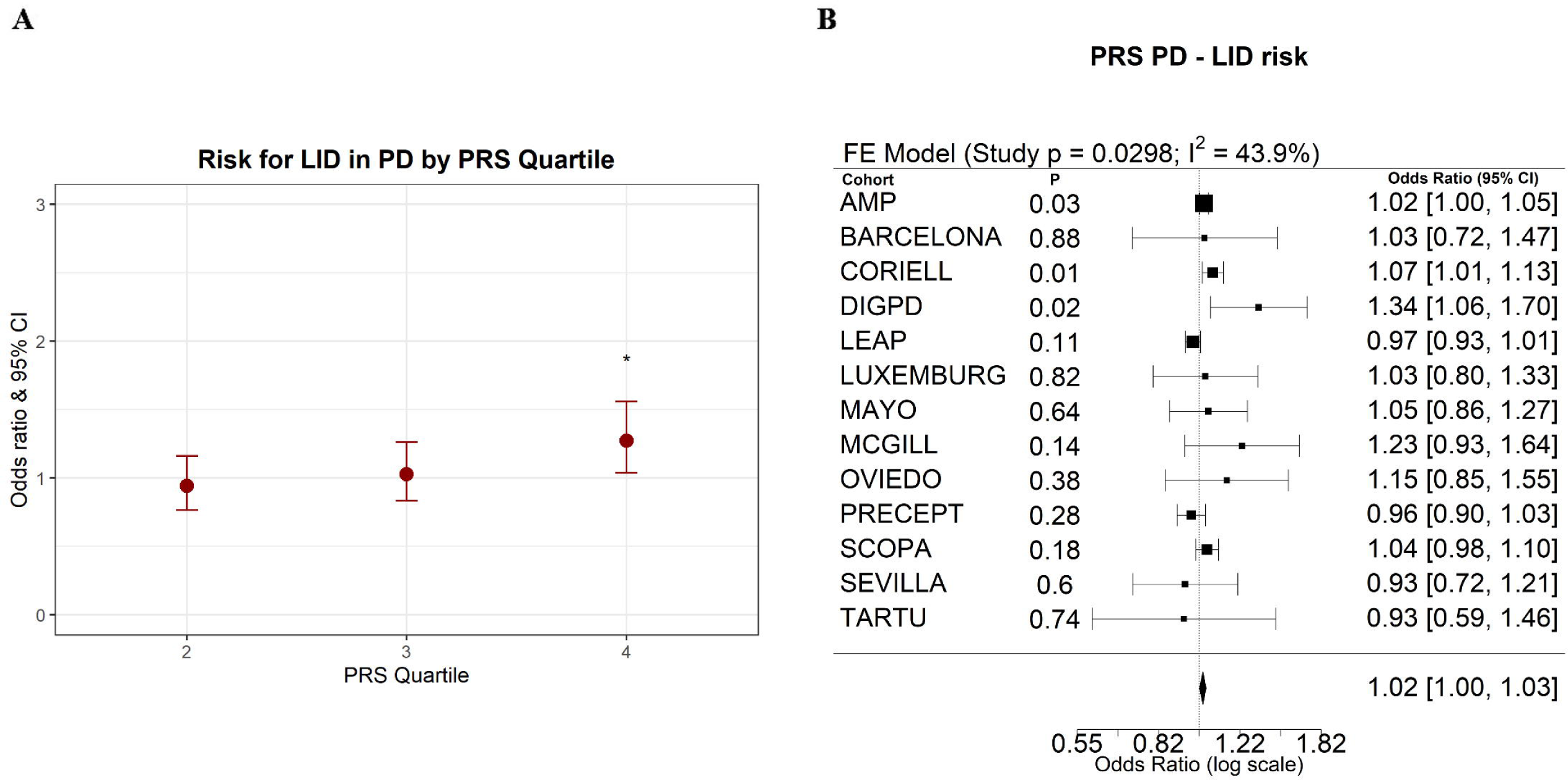
A-B - Logistic regression between PRS aggregating PD risk variants and LID risk. **A.** The plot shows the association between each PRS quartile and LID risk compared with the first quartile, meta-analyzing the results across the cohorts. The Y axis represents the PRS quartile, the X axis the odds ratio (red dot) and 95% confidence interval (red bar). The presence of an asterisk indicates a significant association (*p*<0.05). **B.** The forest plot shows the association between PRS as a continuous variable and LID risk. CI: confidence interval.

### Dopaminergic transmission pathway polygenic effect score is associated with a reduced time to development of LID

Analyses on the dopaminergic transmission pathway PES showed that higher values of PES were associated with a reduced time to development of LID (HR=1.10, 95% CI=1.02-1.18, *p*=0.0088, Fig. 4B). In addition, the third and fourth PES quartile were also associated with a reduced time to development of LID with a more elevated effect size compared to the analyses on PES as a continuous variable (HR_third_quartile=_1.38, 95% CI=1.07-1.79, *p=*0.0128; HR_fourth_quartile=_1.38, 95% CI=1.06-1.78, *p=*0.0147, Fig. 4A, Supplementary Table 10). Logistic regression did not show any statistically significant associations between dopaminergic transmission PES and LID risk (Supplementary Fig. 6 A-B, Supplementary Table 9).

**Fig. 4.**
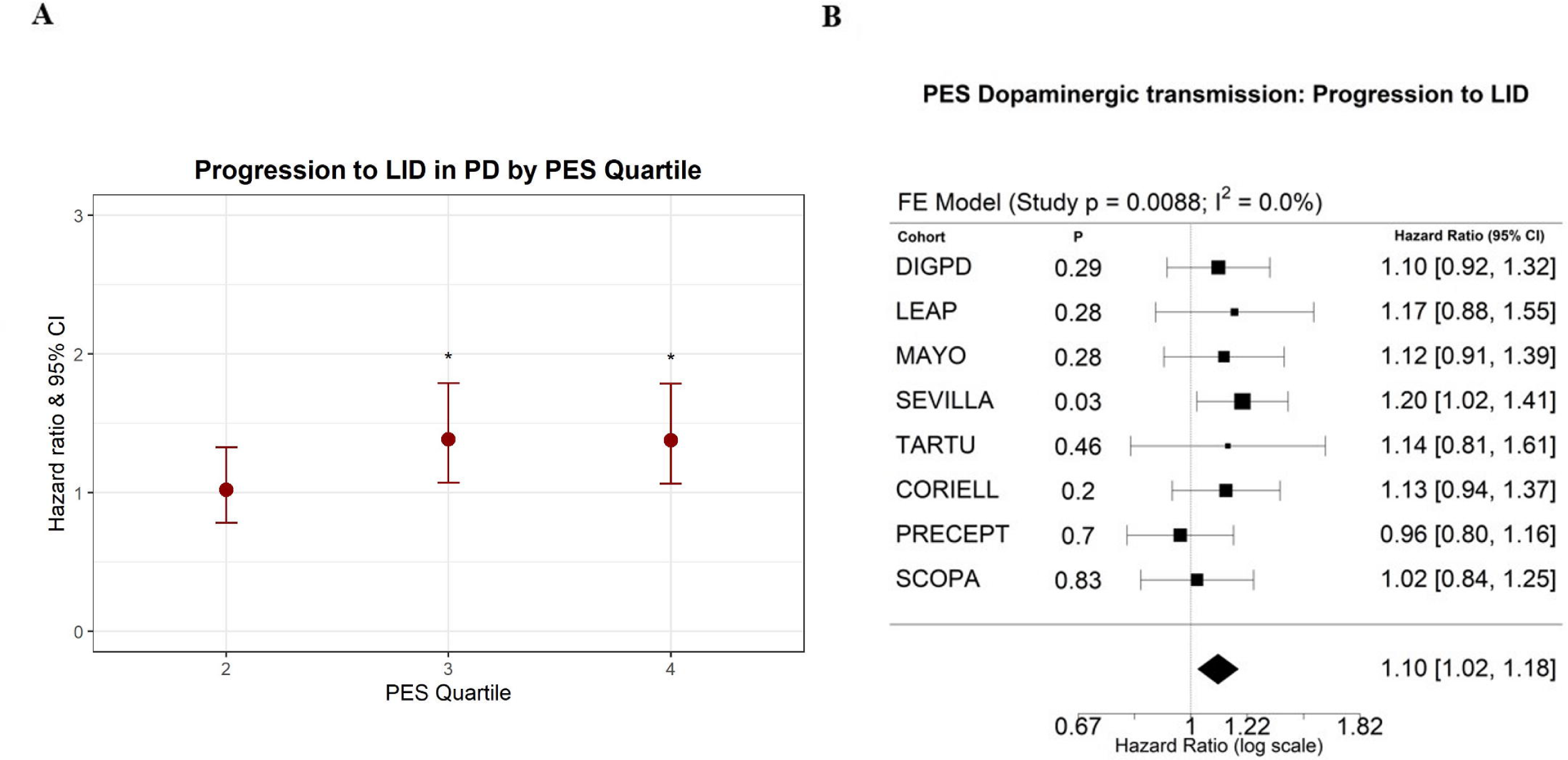
A-B - Cox regression between the dopaminergic transmission pathway PES and time to development of LID. **A.** The plot shows the association between each PES quartile and time to development of LID compared with the first quartile, meta-analyzing the results across the cohorts. The Y axis represents the PRS quartile, the X axis the hazard ratio (red dot) and 95% confidence interval (red bar). **B.** The forest plot shows the association between PES as a continuous variable and time to development of LID.

## Discussion

In this study, we confirmed that *GBA1* variants were associated with increased risk for LID and demonstrated that *LRRK2* variants were associated with a reduced time to development of LID. Additionally, we found that PD PRS was associated with mildly increased risk for LID and that the dopaminergic transmission pathway PES is associated with a reduced time to development of LID.

Albeit some studies found contradictory results on the association between the *GBA1* and *LRRK2* variants and LID,^39–42^ many others have shown that these variants play a role in LID development, ^27–32^ and in this study we also demonstrated that *LRRK2* variants might also affect the time to development of LID. The absence of significant signals in the risk and progression GWAS and, in general, the difficulty finding congruent results between different genetic studies investigating LID, as also reflected by the divergent results between the recent LID progression GWAS (Martinez et al., 2023, MedRxiv) and our study, may be due to the stronger contribution in LID development of environmental factors, especially pharmacologic- (dosage of dopaminergic drugs, use of amantadine) and disease-related factors.^9–13^

The significant association between the two PRS analyses suggests that aggregating multiple common variants that might have a scarce effect on LID individually could contribute to uncovering the overall genetic impact on LID. In particular, the association between the PRS including PD risk variants suggests that patients with a stronger genetic risk profile for PD are also more at risk for LID, a factor to consider for patient counselling and potential clinical trials, although the magnitude of the increased risk was small. We also demonstrated that the dopaminergic synaptic transmission pathway PES was associated with an increased rate of LID development, which is in line with previous pathophysiologic hypotheses^6–8^ and studies suggesting an implication of dopamine pathway genes in the development of LID.^14–18, 21–24^

Unravelling the etiologic bases of LID is crucial to implement a tailored therapy for PD patients taking levodopa, adapting the therapeutic choices, dosage and management depending on the individual risk factors of each patient. Over time, it could be beneficial to define a risk profile accounting for the single genetic and environmental factors associated with LID as well as the cumulative genetic risk provided by the PRS. This might be used to stratify patients for LID prevention clinical trials and lead to a more refined and personalized therapeutic approach for each individual. In addition to the benefits of the current symptomatic therapies, uncovering and confirming genetic factors affecting the risk and time to development of LID could also have important implications for targeted therapies. In particular, *GBA1* and *LRRK2* pathways are already candidate targets for newly developing drugs in clinical trials.^49^ A LRRK2 inhibitor, BIIB122/DNL151, reached already experimental phase 3 (https://www.denalitherapeutics.com, 2021).^57^ In addition, Ambroxol, a pharmacological chaperone for GCase capable of increasing its enzymatic levels, completed phase 2 and LTI-291, an activator of GCase, reached phase 1B.^58–60^ As these drugs would likely be used in conjunction with symptomatic therapies, knowing that these pathways can be targeted to reduce the risk or delay the time of LID development could considerably improve the compliance and quality of life of PD patients taking dopaminergic treatments.

The current study has several limitations. First, the subjects were all of European ancestry and therefore the results in other populations might be different. Despite an overall large sample size, most of the individual cohorts included a limited number of participants, especially those having longitudinal data necessary for Cox regression, this impacted the power of the study and could have contributed to the lack of association in the GWAS. Some studies suggested that LID is affected more by the disease duration than by the therapy duration,^61^ on this line PD AAO would represent a better baseline than levodopa initiation for the time to LID onset. However, this parameter was chosen in accordance with what was previously done with LID GWAS^50^ and accounting for the recall bias that PD AAO suffers from, compared to levodopa initiation which represents a report made by the physicians. In addition, understanding the genetic basis of the time to LID from levodopa initiation can be of considerable relevance for patient counselling at the time of treatment administration. Finally, we also accounted for the disease duration in each of our analyses with appropriate adjustments. Another limitation of this study was that not all the cohorts had the same amount of data available, which limited in part the design of the analytical model.

In conclusion, in the current study we demonstrated that PD risk variants and the dopaminergic transmission PRS are associated with increased risk of LID/time to development of LID. A better understanding of the role of genetics in LID development could reduce the impact of this adverse effect and enhance therapeutic management in PD.

## Disclosures

### Financial Disclosures for the previous 12 months

The authors declare that there are no additional disclosures to report.

## Acknowledgements

We wholeheartedly thank the participants in this study. We would like to thank the research participants and all members of IPDGC for making this work possible. The AMP-PD cohort data used in this study included the Fox Investigation for New Discovery of Biomarkers (BioFIND), the Harvard Biomarker Study (HBS), the Parkinson’s Disease Biomarkers Program (PDBP) and the Parkinson’s Progression Markers Initiative (PPMI) cohorts. For up-to-date information on the study, visit https://www.amp-pd.org. AMP PD – a public-private partnership – is managed by the FNIH and funded by Celgene, GSK, the Michael J. Fox Foundation for Parkinson’s Research, the National Institute of Neurological Disorders and Stroke, Pfizer, Sanofi, and Verily. BioFIND is sponsored by The Michael J. Fox Foundation for Parkinson’s Research (MJFF) with support from the National Institute for Neurological Disorders and Stroke (NINDS). The BioFIND Investigators have not participated in reviewing the data analysis or content of the manuscript. For up-to-date information on the study, visit https://www.michaeljfox.org/news/biofind. The Harvard Biomarker Study (HBS) is a collaboration of HBS investigators [full list of HBS investigators found at https://www.bwhparkinsoncenter.org/biobank/ and funded through philanthropy and NIH and Non-NIH funding sources. The HBS Investigators have not participated in reviewing the data analysis or content of the manuscript. The Parkinson’s Disease Biomarker Program (PDBP) consortium is supported by the National Institute of Neurological Disorders and Stroke (NINDS) at the National Institutes of Health. A full list of PDBP investigators can be found at https://pdbp.ninds.nih.gov/policy. The PDBP investigators have not participated in reviewing the data analysis or content of the manuscript. PPMI is sponsored by The Michael J. Fox Foundation for Parkinson’s Research and supported by a consortium of scientific partners: [list of the full names of all of the PPMI funding partners can be found at https://www.ppmi-info.org/about-ppmi/who-we-are/study-sponsors]. The PPMI investigators have not participated in reviewing the data analysis or content of the manuscript. For up-to-date information on the study, visit www.ppmi-info.org. This research was supported in part by the Intramural Research Program of the NIH, National Institute on Aging (NIA), National Institutes of Health, Department of Health and Human Services; project number ZO1 AG000535 and ZIA AG000949, as well as the National Institute of Neurological Disorders and Stroke (NINDS, program # ZIANS003154).

We would like to thank all participants of the Luxembourg Parkinson’s Study for their important support of our research. Data used for the Luxemburg cohort in the preparation of this manuscript were obtained from the National Centre of Excellence in Research on Parkinson’s Disease (NCER-PD). We acknowledge the joint effort of the NCER-PD Consortium members from the partner institutions Luxembourg Centre for Systems Biomedicine, Luxembourg Institute of Health, Centre Hospitalier de Luxembourg, and Laboratoire National de Santé generally contributing to the Luxembourg Parkinson’s Study as listed below: Geeta ACHARYA 2, Gloria AGUAYO 2, Myriam ALEXANDRE 2, Muhammad ALI 1, Wim AMMERLANN 2, Giuseppe ARENA 1, Rudi BALLING 1, Michele BASSIS 1, Katy BEAUMONT 2, Regina BECKER 1, Camille BELLORA 2, Guy BERCHEM 3, Daniela BERG 11, Alexandre BISDORFF 5, Ibrahim BOUSSAAD 1, Kathrin BROCKMANN 11, Jessica CALMES 2, Lorieza CASTILLO 2, Gessica CONTESOTTO 2, Nico DIEDERICH 3, Rene DONDELINGER 5, Daniela ESTEVES 2, Guy FAGHERAZZI 2, Jean-Yves FERRAND 2, Manon GANTENBEIN 2, Thomas GASSER 11, Piotr GAWRON 1, Soumyabrata GHOSH 1, Marijus GIRAITIS 2,3, Enrico GLAAB 1, Elisa GÓMEZ DE LOPE 1, Jérôme GRAAS 2, Mariella GRAZIANO 17, Valentin GROUES 1, Anne GRÜNEWALD 1, Wei GU 1, Gaël HAMMOT 2, Anne-Marie HANFF 2, Linda HANSEN 1,3, Michael HENEKA 1, Estelle HENRY 2, Sylvia HERBRINK 6, Sascha HERZINGER 1, Michael HEYMANN 2, Michele HU 8, Alexander HUNDT 2, Nadine JACOBY 18, Jacek JAROSLAW LEBIODA 1, Yohan JAROZ 1, Sonja JÓNSDÓTTIR 2, Quentin KLOPFENSTEIN 1, Jochen KLUCKEN 1,2,3, Rejko KRÜGER 1,2,3, Pauline LAMBERT 2, Zied LANDOULSI 1, Roseline LENTZ 7, Inga LIEPELT 11, Robert LISZKA 14, Laura LONGHINO 3, Victoria LORENTZ 2, Paula Cristina LUPU 2, Tainá M. MARQUES 1, Clare MACKAY 10, Walter MAETZLER 15, Katrin MARCUS 13, Guilherme MARQUES 2, Patricia MARTINS CONDE 1, Patrick MAY 1, Deborah MCINTYRE 2, Chouaib MEDIOUNI 2, Francoise MEISCH 1, Myriam MENSTER 2, Maura MINELLI 2, Michel MITTELBRONN 1,4, Brit MOLLENHAUER 12, Friedrich MÜHLSCHLEGEL 4, Romain NATI 3, Ulf NEHRBASS 2, Sarah NICKELS 1, Beatrice NICOLAI 3, Jean-Paul NICOLAY 19, Fozia NOOR 2, Marek OSTASZEWSKI 1, Clarissa P. C. GOMES 1, Sinthuja PACHCHEK 1, Claire PAULY 1,3, Laure PAULY 1, Lukas PAVELKA 1,3, Magali PERQUIN 2, Rosalina RAMOS LIMA 2, Armin RAUSCHENBERGER 1, Rajesh RAWAL 1, Dheeraj REDDY BOBBILI 1, Kirsten ROOMP 1, Eduardo ROSALES 2, Isabel ROSETY 1, Estelle SANDT 2, Stefano SAPIENZA 1, Venkata SATAGOPAM 1, Margaux SCHMITT 2, Sabine SCHMITZ 1, Reinhard SCHNEIDER 1, Jens SCHWAMBORN 1, Amir SHARIFY 2, Ekaterina SOBOLEVA 1, Kate SOKOLOWSKA 2, Hermann THIEN 2, Elodie THIRY 3, Rebecca TING JIIN LOO 1, Christophe TREFOIS 1, Johanna TROUET 2, Olena TSURKALENKO 2, Michel VAILLANT 2, Mesele VALENTI 2, Gilles VAN CUTSEM 1,3, Carlos VEGA 1, Liliana VILAS BOAS 3, Maharshi VYAS 1, Richard WADE-MARTINS 9, Paul WILMES 1, Evi WOLLSCHEID-LENGELING 1, Gelani ZELIMKHANOV 3

1 Luxembourg Centre for Systems Biomedicine, University of Luxembourg, Esch-sur-Alzette, Luxembourg

2 Luxembourg Institute of Health, Strassen, Luxembourg

3 Centre Hospitalier de Luxembourg, Strassen, Luxembourg

4 Laboratoire National de Santé, Dudelange, Luxembourg

5 Centre Hospitalier Emile Mayrisch, Esch-sur-Alzette, Luxembourg

6 Centre Hospitalier du Nord, Ettelbrück, Luxembourg

7 Parkinson Luxembourg Association, Leudelange, Luxembourg

8 Oxford Parkinson’s Disease Centre, Nuffield Department of Clinical Neurosciences, University of Oxford, Oxford, UK

9 Oxford Parkinson’s Disease Centre, Department of Physiology, Anatomy and Genetics, University of Oxford, Oxford, UK

10 Oxford Centre for Human Brain Activity, Wellcome Centre for Integrative Neuroimaging, Department of Psychiatry, University of Oxford, Oxford, UK

11 Center of Neurology and Hertie Institute for Clinical Brain Research, Department of Neurodegenerative Diseases, University Hospital Tübingen, Tübingen, Germany

12 Paracelsus-Elena-Klinik, Kassel, Germany

13 Ruhr-University of Bochum, Bochum, Germany

14 Westpfalz-Klinikum GmbH, Kaiserslautern, Germany

15 Department of Neurology, University Medical Center Schleswig-Holstein, Kiel, Germany

16 Department of Neurology Philipps, University Marburg, Marburg, Germany

17 Association of Physiotherapists in Parkinson’s Disease Europe, Esch-sur-Alzette, Luxembourg

18 Private practice, Ettelbruck, Luxembourg

19 Private practice, Luxembourg, Luxembourg

PreCEPT and PostCEPT were funded by NINDS 5U01NS050095- 05, Department of Defense Neurotoxin Exposure Treatment Parkinson’sResearchProgram(Grant Number: W23RRYX7022N606), the Michael J Fox Founda- tion for Parkinson’s Research, Parkinson’s Disease Foundation, Lundbeck Pharmaceuticals. Cephalon Inc, Lundbeck Inc, John Blume Foundation, Smart Family Foundation, RJG Founda- tion, Kinetics Foundation, National Parkinson Foundation, Amarin Neuroscience LTD, CHDI Foundation Inc, NIH (NHGRI and NINDS), and Columbia Parkinson’sDisease Research Center. ZGO is supported by the Fonds de recherche du Québec - Santé (FRQS) Chercheurs-boursiers award, and is a William Dawson Scholar. YLS is supported by the HBHL Graduate student fellowship. ProPARK is funded by the Alkemade-Keuls Foundation, Stichting Parkinson Fonds, Parkinson Vereniging, and The Netherlands Organization for Health Research and Development; Udall is supported by the NINDS. This work at the Mayo Clinic Florida was supported by: the Haworth Family Professorship in Neurodegenerative Diseases fund, and The Albertson Parkinson’s Research Foundation.

## Author roles

1. Research project: A. Conception, B. Organization, C. Execution;

2. Cohort generation: A. Cohort recruitment, B. Sample processing, C. Data generation;

3. Statistical Analysis: A. Design, B. Execution, C. Review and Critique;

4. Manuscript Preparation: A. Writing of the first draft, B. Review and Critique

YLS: 1A, 1B, 1C, 3A, 3B, 4A, 4B

SBC: 2C, 4B

BF: 3B, 4B

RMADB: 1A, 4B

MTPT: 2C, 4B

RDB: 2C, 4B

PGG: 2C, 4B

PM: 1A, 4B

JF: 3B, 4B

PT: 2C, 4B

JCC: 1A, 4B

LP: 2C, 4B

TM: 2C, 4B

CG: 2C, 4B

AK: 2C, 4B

PM: 2B, 4B

RK: 1A, 4B

LMM: 2C, 4B

ZKW: 1B, 4B

RJU: 1B, 4B

OR: 1A, 4B

PH: 1B, 4B

JH: 1B, 4B

DKS: 1B, 4B

SE: 1B, 4B

LK: 3C, 4B

EY: 3C, 4B

UR: 2B, 4B

JAR: 2B, 4B

KF: 2B, 4B

FA: 2B, 4B

MM: 2C, 4B

IAF: 2C, 4B

PP: 1C, 4B

SK: 1C, 2C, 4B

HI: 1C, 3A, 3B, 3C, 4B

ZGO: 1A, 1B, 2A, 3A, 3C, 4B

## Ethical Compliance Statement

IRB Study Number A11-M60-21A (21-11-023) was reviewed and approved by the Research Ethics Offices (REOs). Informed written patient consent was provided in each center before the inclusion of each in the study. We confirm that we have read the Journal’s position on issues involved in ethical publication and affirm that this work is consistent with those guidelines

## Data availability

The LID GWAS summary statistics are publicly available on GWAS catalog (https://www.ebi.ac.uk/gwas/). All codes used for the analyses are available at https://github.com/gan-orlab.

