## Supplementary Figures for "Dopamine pathway and Parkinson’s risk variants are associated with levodopa-induced dyskinesia"

**Supplementary Fig. 1 – Q-Q plot of LID risk GWAS using logistic regression**

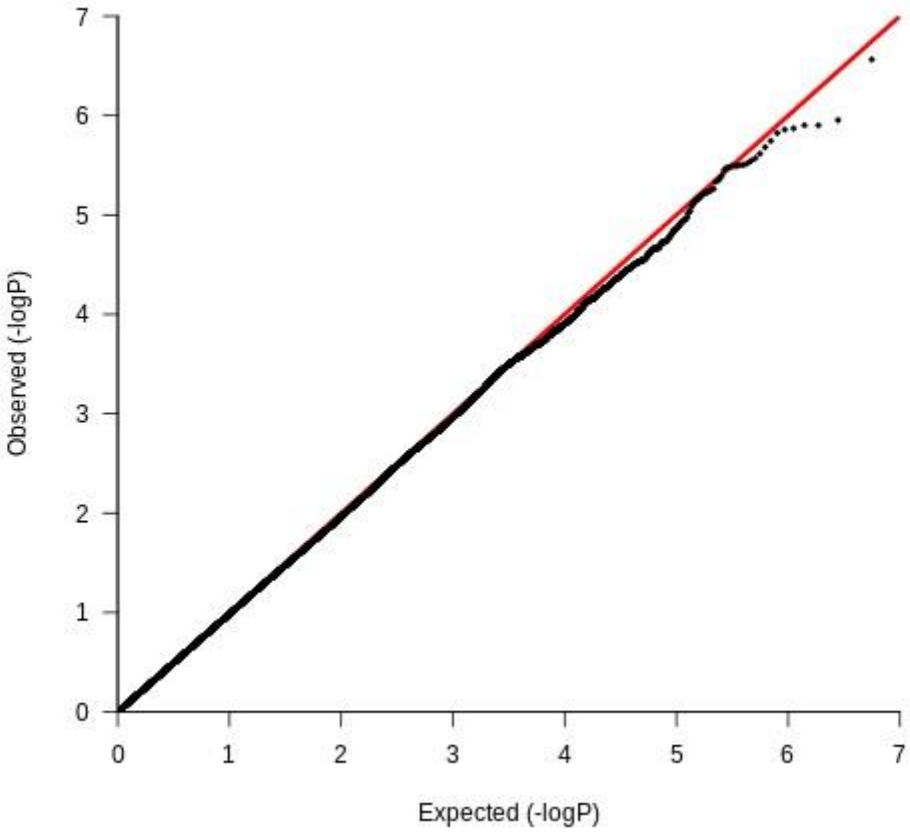

**Supplementary Fig. 2 - Q-Q plot of progression to LID GWAS using Cox regression**

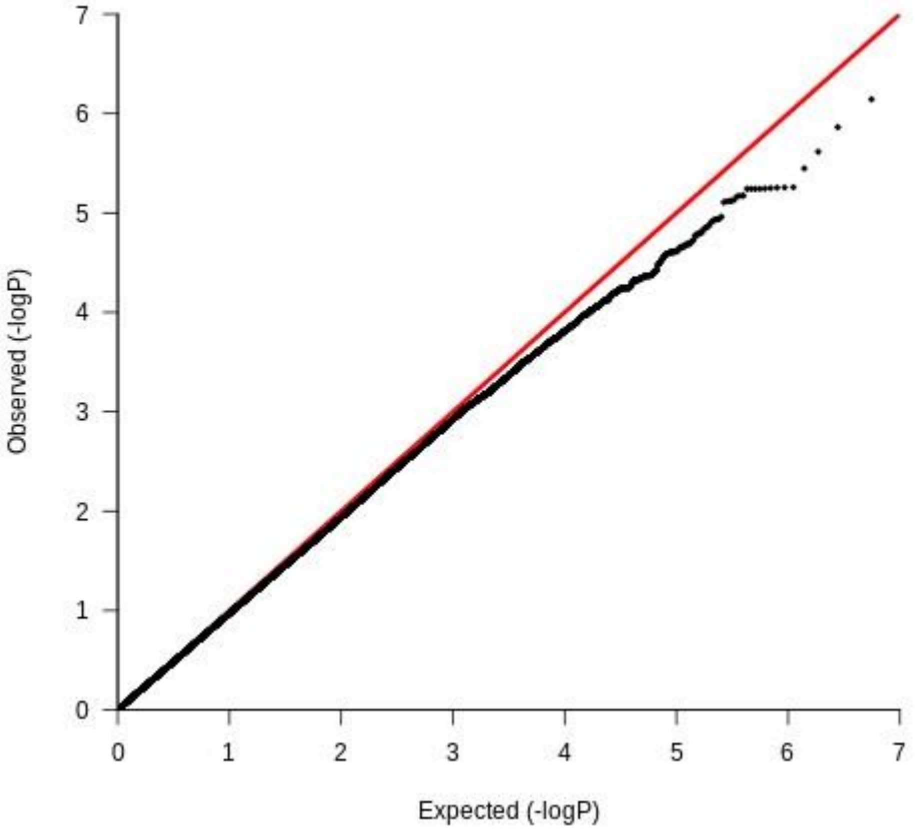

**Supplementary Fig. 3 – Manhattan plot of LID risk GWAS**

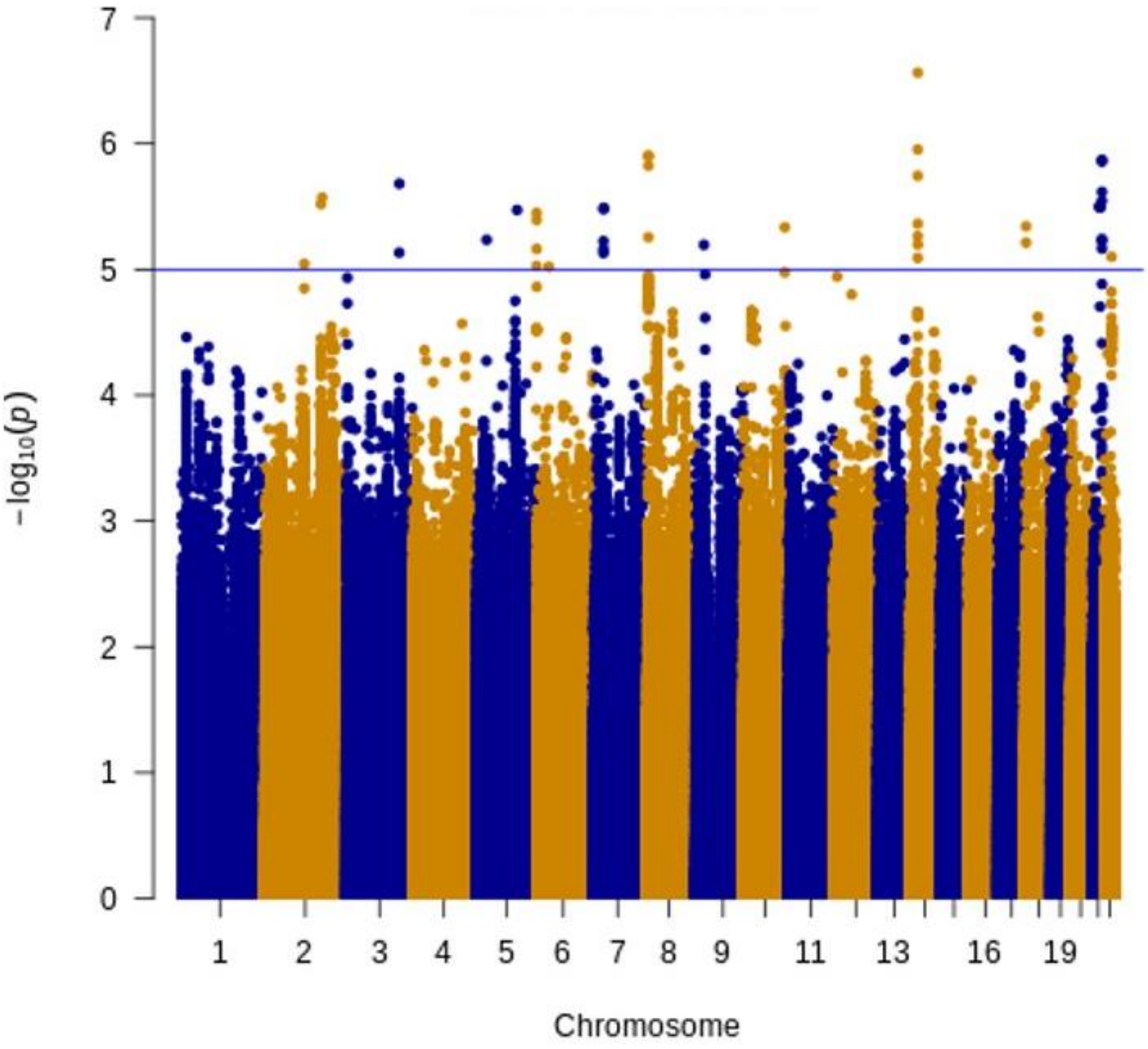

**Supplementary Fig. 4 – Manhattan plot of progression to LID GWAS**

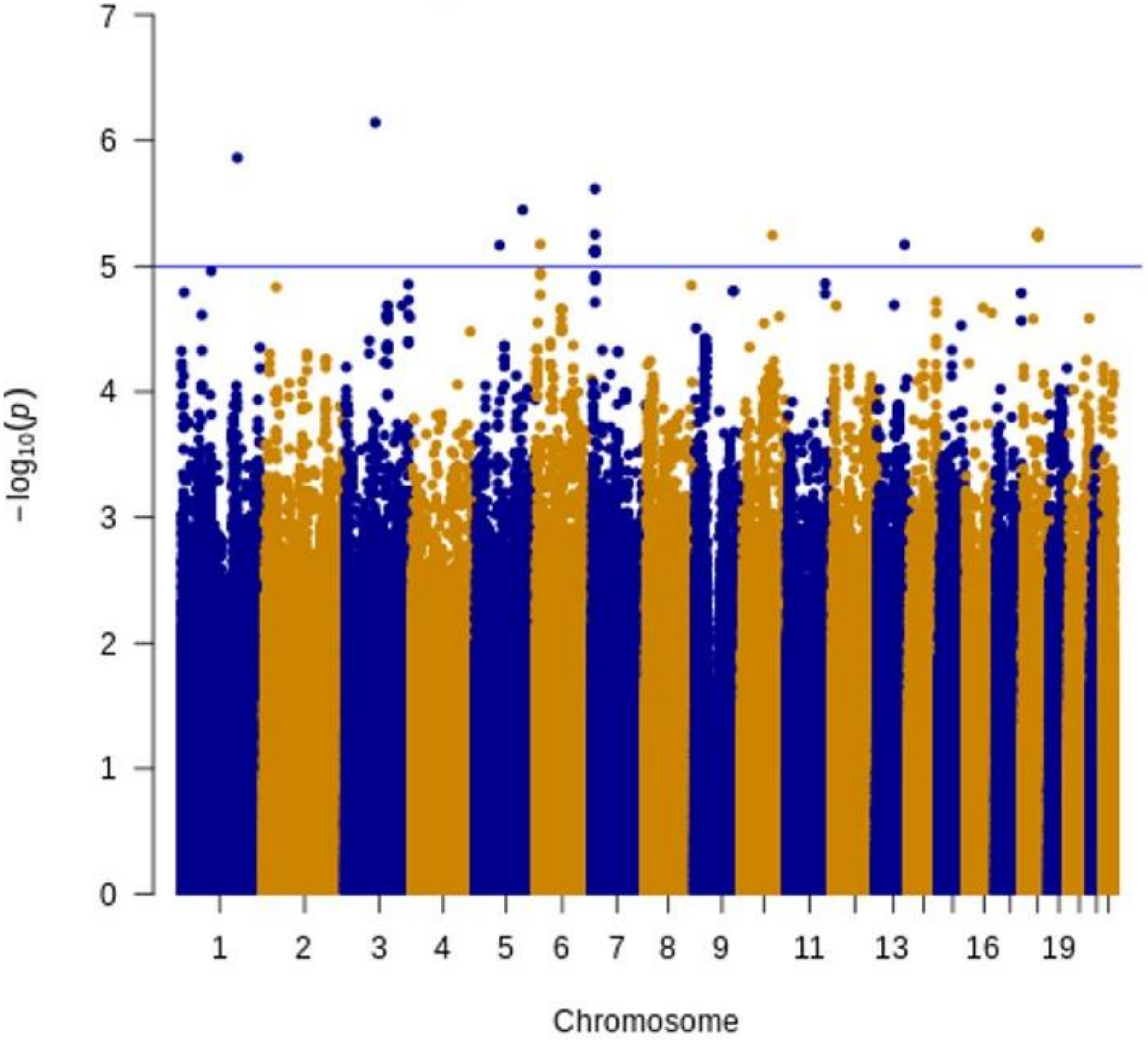

**Supplementary Fig. 5 - A-B - Cox regression between PRS including PD risk variants and progression to LID**

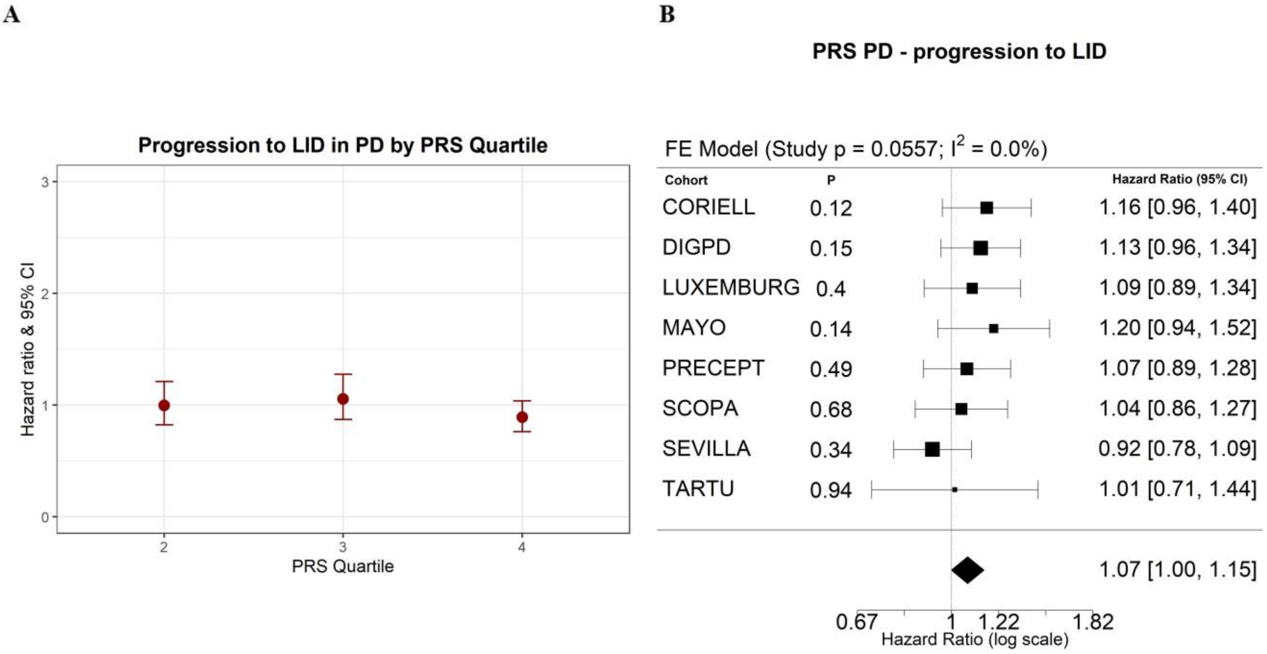

**Supplementary Fig. 6 A-B - Logistic regression between the dopaminergic transmission pathway PES and LID risk**

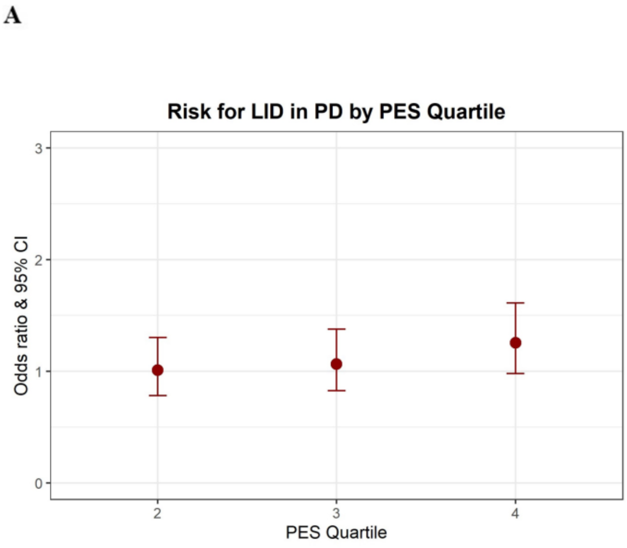


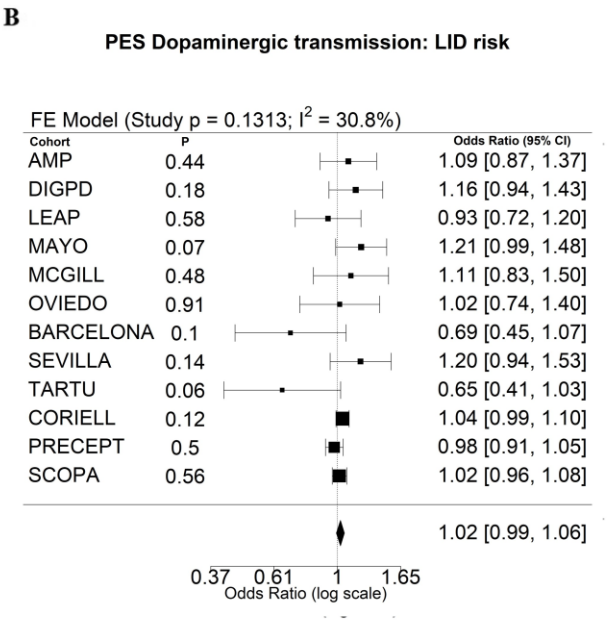
